# Backward and Hopf bifurcation analysis of an SEIRS COVID-19 epidemic model with saturated incidence and saturated treatment response

**DOI:** 10.1101/2020.08.28.20183723

**Authors:** David A. Oluyori, Ángel G. C. Pérez, Victor A. Okhuese, Muhammad Akram

**Author notes:** Corresponding author. E-mail addresses (D. A. Oluyori), (A. G. C. Pérez), (V. A. Okhuese), (M. Akram).

## Abstract

In this work, we further the investigation of an SEIRS model to study the dynamics of the Coronavirus Disease 2019 pandemic. We derive the basic reproduction number *R*_0_ and study the local stability of the disease-free and endemic states. Since the condition *R*_0_ < 1 for our model does not determine if the disease will die out, we consider the backward bifurcation and Hopf bifurcation to understand the dynamics of the disease at the occurrence of a second wave and the kind of treatment measures needed to curtail it. Our results show that the limited availability of medical resources favours the emergence of complex dynamics that complicates the control of the outbreak.

## 1 Introduction

Since the first cases of Coronavirus Disease 2019 (COVID-19) were reported in Wuhan, China in December 2019, the number of cases of this disease has increased exponentially and the pandemic has become a global threat. The outbreak of COVID-19 has spread all over the world and has been declared a public health emergency by the World Health Organization (WHO). Many researchers have proposed mathematical models based on systems of differential equations to describe the dynamics of the COVID-19 outbreak as time progresses, and these models have been proved useful to investigate the effect of applying different strategies to contain the epidemic.

In the paper [24], the authors discussed the global analysis of an SEIRS model for COVID-19 with saturated incidence and treatment response where, among other results, they derived the basic reproduction number and established the local and global stabilities of the disease-free and endemic states, thus concluding that the effectiveness of the treatment response applied determines whether an endemic situation is imminent within the population. The SEIRS model proposed in [24] is an extension the SEIRUS model studied in [28, 27] (where *S* represents the susceptible class, *E* is the exposed class in the latent period, *I* is the infectious class, *R* is the recovered class and *U* is the undetectable class), obtained as a result of collapsing the *U* class due to the fact that the undetected class and the recovered class can only be certified COVID-19 free when they test negative twice. The SEIRUS model as conceived by the Centre for Disease Control in 2017 was built on the premise that on recovery there is no reinfection. Currently, there is no definite answer to the question whether people who recover from COVID-19 can be reinfected with the virus [20]. It is not clear whether some patients who have recovered and later tested positive again have truly been reinfected, or, at the time of their “recovery”, they still had low levels of the virus in their systems [20, 21].

There are growing concerns from trend analysis of COVID-19 that endemic situations may be imminent in some parts of the world such that when the current disease trend is long forgotten, the disease may become endemic in some parts as in the case of Lassa Fever in Nigeria [14], Ebola in D.R. Congo [6] to mention a few.

Some studies [15, 18, 29] have shown that the dynamics of the model proposed are determined by the disease’s basic reproduction number, *R*_0_. The fact which is generally known to epidemiologists that if *R*_0_ < 1, the disease can be eliminated from the community, whereas an epidemic occurs when *R*_0_ > 1 [8]. Meanwhile, other studies such as Alexander et al. [4] and Arino et al. [5] established that the criterion for *R*_0_ < 1 is not always sufficient to control the spread of the disease, a phenomenon known as *backward bifurcation*. Mathematically, when a backward bifurcation occurs, there are at least three equilibria for a certain range of parameters with *R*_0_ < 1: the stable disease-free equilibrium, a large stable endemic equilibrium and a small unstable endemic equilibrium which acts as a boundary between the basins of attraction for the two stable equilibria. In some cases, a backward bifurcation leading to bistability can occur, which makes the disease endemic in the population given a sufficiently large initial outbreak. Several examples of this bistable behaviour have been found in mathematical models for COVID-19, such as [21, 3, 22, 17]. These phenomena pose significant epidemiological consequences for disease management since their existence implies that the basic reproduction number of the disease should be reduced to a value much lower than one to ensure the eradication of the epidemic.

A common assumption in classical epidemic models is that the rate of treatment against the disease is directly proportional to the number of infective individuals. However, in the case of COVID-19, the availability of medical resources such as ventilators, hospital beds and trained medical personnel is too limited compared to the increasing number of infected cases, which has inflicted a great pressure on the healthcare systems around the world. Hence, when developing mathematical models for this disease, it would be more adequate to consider a saturated treatment rate, which increases more slowly when the size of infected population becomes too large.

## 2 Model description

In this paper, we propose to study the following model:

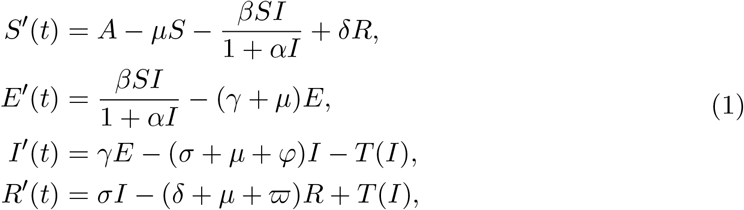

with the total population being

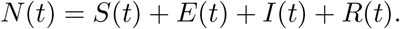

Here, *S*(*t*) is the number of susceptible individuals, *E*(*t*) is the number of exposed individuals, *I*(*t*) is the number of infectious individuals, and *R*(*t*) is the number of individuals that are quarantined and expecting recovery or have recovered from the infection.

The parameter *A* is the recruitment rate of the population, *μ* is the natural death rate of the population per time unit, *α* is the saturation parameter that measures the inhibitory effect, *β* is the rate of transmission, *γ* is the rate of developing infection after being exposed, *σ* is the natural recovery rate, *δ* is the proportion of the removed population that becomes susceptible again, *φ* is the disease-induced death rate of the infected population not quarantined, *ϖ* is the disease-induced death rate of quarantined infected population, *βSI/*(1 + *αI*) is the saturated incidence rate, 1*/*(1 + *αI*) is the inhibitory factor and *T* (*I*) is the saturated treatment response defined as

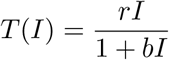

(here if *b* = 0, the treatment becomes bilinear, and if *r* = 0, the treatment is null). All parameters are assumed positive, except *δ*, which is non-negative. Notice that *δ* = 0 represents the case when reinfection is not possible, that is, recovery from the disease gives permanent immunity. The flow diagram of the model can be seen in Figure 1.

**Figure 1:**
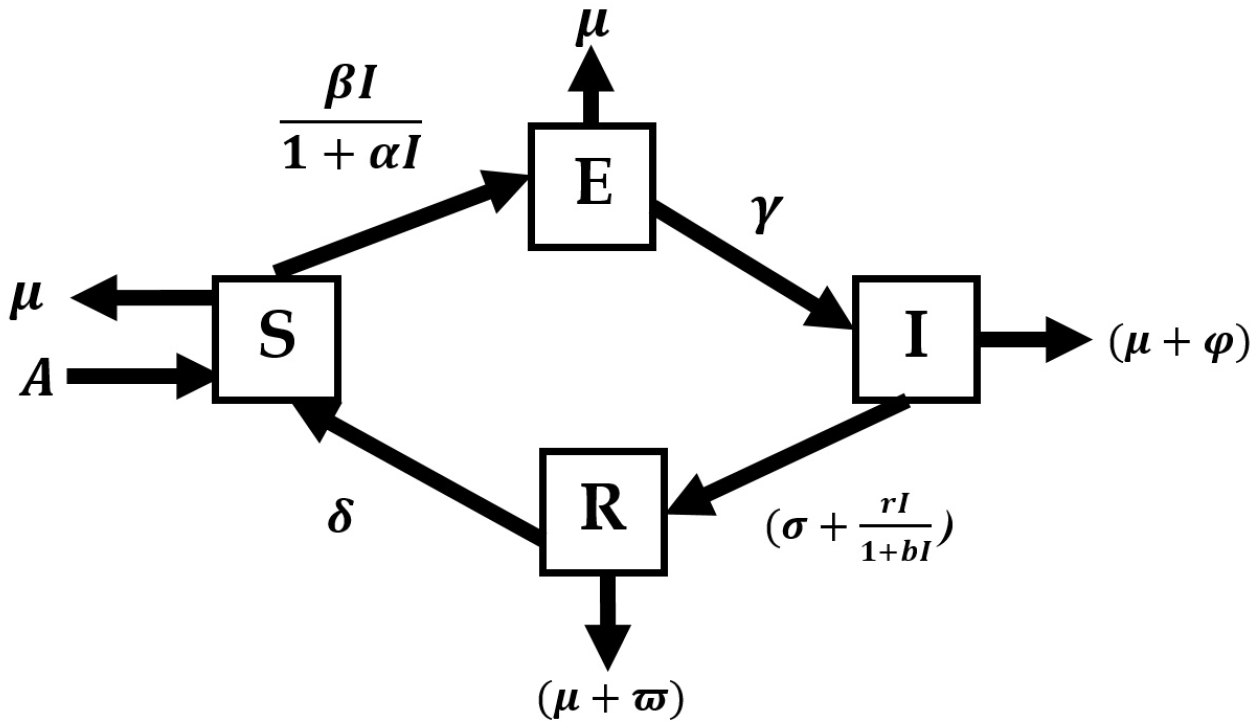
Schematic diagram of the model.

The SEIRS model (1) is based on the one studied in [24] but replacing the piecewise linear treatment function by a saturated function. We can see that model (1) is similar to Agrawal et al.’s model in [2]. Here, we will extend the research done in [24] and [2] by studying the backward and Hopf bifurcation dynamics of model (1).

The rest of this paper is structured as follows. In Section 3, we compute the basic reproduction number of our model. In Section 4, we determine conditions for the existence of endemic equilibria. We study the local stability of equilibria in Section 5. We provide conditions for the occurrence of backward bifurcation in Section 6 and for Hopf bifurcation in Section 7. In Section 8, we fit the parameter values to the reported data of the COVID-19 pandemic and perform some numerical simulations. Finally, we discuss our results and provide some conclusions in Section 9.

## 3 Basic reproduction number

Considering the feasible region of the system (1) as

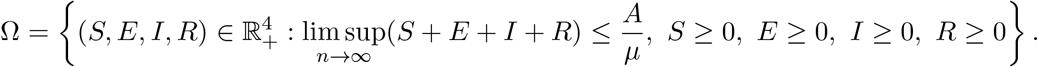

It follows that the region Ω is positively invariant with respect to the system (1), which implies that the *ω*-limit sets of all solutions of the model in 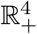 are contained in Ω for all time and those outside Ω are eventually attracted to that region. Hence, the system (1) is epidemiologically well posed.

Now, we will find the basic reproduction number *R*_0_ of the system (1) by obtaining the Jacobian of the system and using the next generation matrix due to van den Driessche and Watmough [13]. The Jacobian matrix evaluated at en equilibrium (*S*^*\**^, *E*^*\**^, *I*^*\**^, *R*^*\**^) is

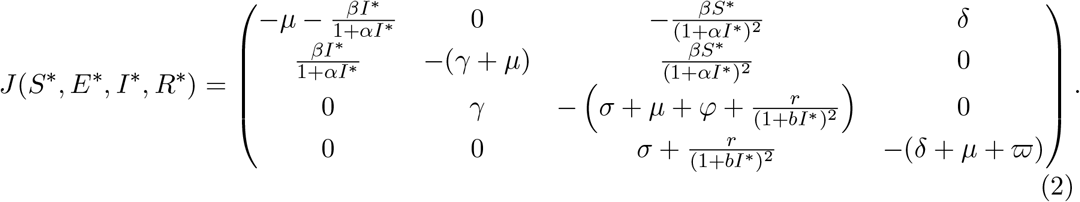

We derive the disease-free equilibrium of system (1) with *I* = 0 and *S* = *A/μ* as the Jacobian of the system in (2) reduces to

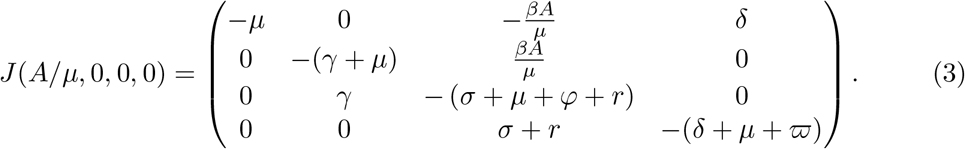

Using the next generation matrix, it is clear that the reproduction number *R*_0_ is the spectral radius of the next generation matrix derived from the exposed and infected class, i.e.,

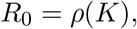

where *ρ*(*K*) is the spectral radius of the operator *K* and *K* = *FV* ^−1^ is the next generation matrix. *F* is derived from the exposed and infected class and *V* are the remaining terms after *F* is taken.

Thus,

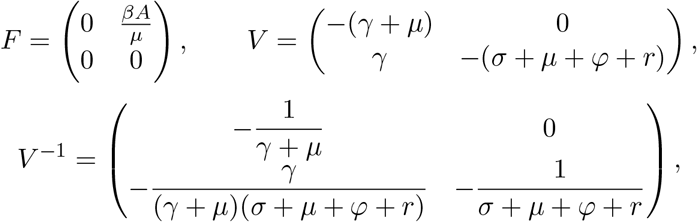

so

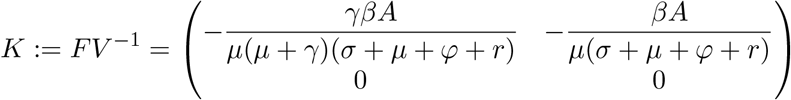

is the next generation matrix of the system (1). The basic reproduction number *R*_0_ of system (1) is given by the spectral radius of *K*. Hence,

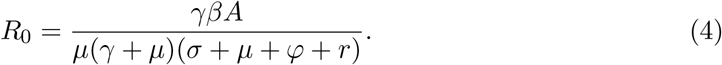

## 4 Equilibria of the model

The equilibria of model (1) are given by the system of equations

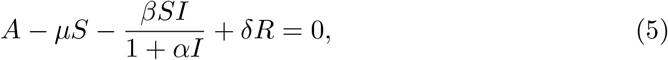

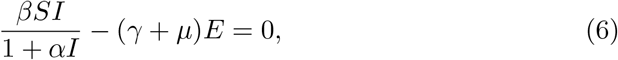

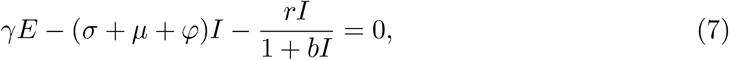

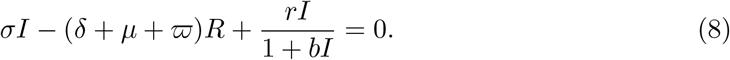

If we substitute *I* = 0 in the above system, we obtain the disease-free equilibrium of the model, given by 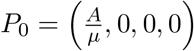.

Regarding the existence of endemic equilibria, we can establish the following result.

### Theorem 1.

*Let*

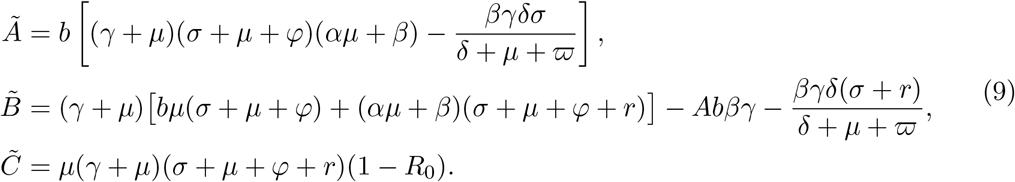

*Then, model* (1) *has:*

i. *A unique endemic equilibrium if R*_0_ > 1;
ii. *A unique endemic equilibrium if* 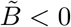, *and R*_0_ = 1 *or* 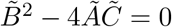;
iii. *Two endemic equilibria if* 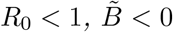 *and* 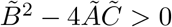;
iv. *No endemic equilibrium otherwise*.

*Proof*. See Appendix A. □

By this, it is clear from Theorem 1 case (i) that the model has a unique equilibrium whenever *R*_0_ > 1. We also see from case (iii) that there exists the possibility of backward bifurcation, which is when the locally asymptotically stable disease-free equilibrium coexists with a locally endemic equilibrium.

Notice that, when *b* = 0, the coefficient 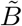 reduces to

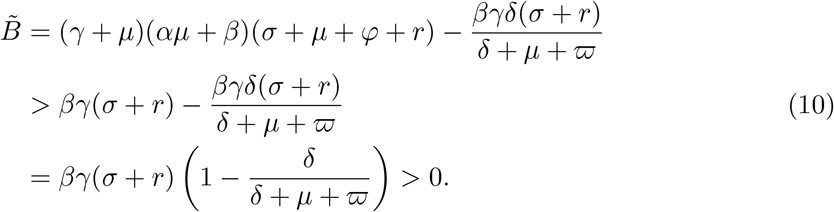

This shows that 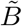 will always be positive for *b* > 0 sufficiently small. Thus, it is noted from Theorem 1 that the backward bifurcation phenomenon will not take place if *b* is close to 0, which occurs when the saturation effect of the non-linear treatment rate *rI/*(1 + *bI*) is reduced.

## 5 Local stability analysis

In this section, we will study the local stability of the disease-free and endemic equilibria of model (1).

### 5.1 Local stability of the disease-free equilibrium

#### Theorem 2.

*The disease-free equilibrium (P*_0_*) is:*

a. *Locally asymptotically stable if R*_0_ < 1.
b. *Unstable if R*_0_ > 1.

*Proof*. The Jacobian matrix of the system at *P*_0_ is given by (3), so the characteristic equation is

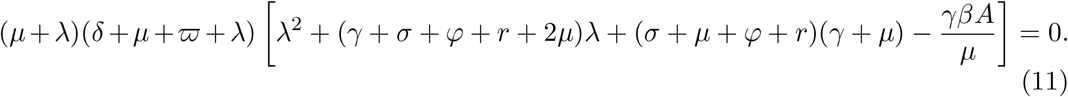

It is clear that *λ*_1_ = −*μ* and *λ*_2_ = −(*δ* + *μ* + *ϖ*) are two roots of the characteristic equation. The other roots are determined by the equation

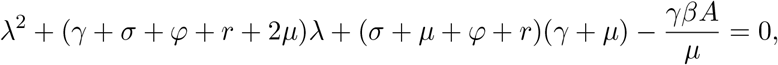

which has negative roots if and only of 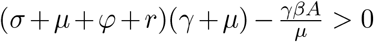, which implies that the reproduction number *R*_0_ is less than one. This implies that the disease-free equilibrium *P*_0_ is locally asymptotically stable when *R*_0_ < 1 and unstable when *R*_0_ > 1. □

Thus, having establish the threshold quantity *R*_0_ in the system (1), which measures the average number of infections generated by a single infected individual in a completely susceptible population, Theorem 2 states that when *R*_0_ < 1, the introduction of a small number of infected individuals into the community would not lead to large outbreak and the disease dies out in time but when *R*_0_ > 1, the disease persists. However, the study of backward bifurcation shows that the disease may persist even when the reproduction number is less than unity (*R*_0_ < 1), as we will study in Section 6.

### 5.2 Local stability of the endemic equilibrium

#### Theorem 3.

*Let P*^*\**^(*S*^*\**^, *E*^*\**^, *I*^*\**^, *R*^*\**^) *be an endemic equilibrium of* (1). *Then P*^*\**^ *is locally asymptotically stable if and only if*

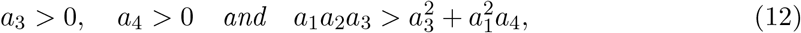

*where*

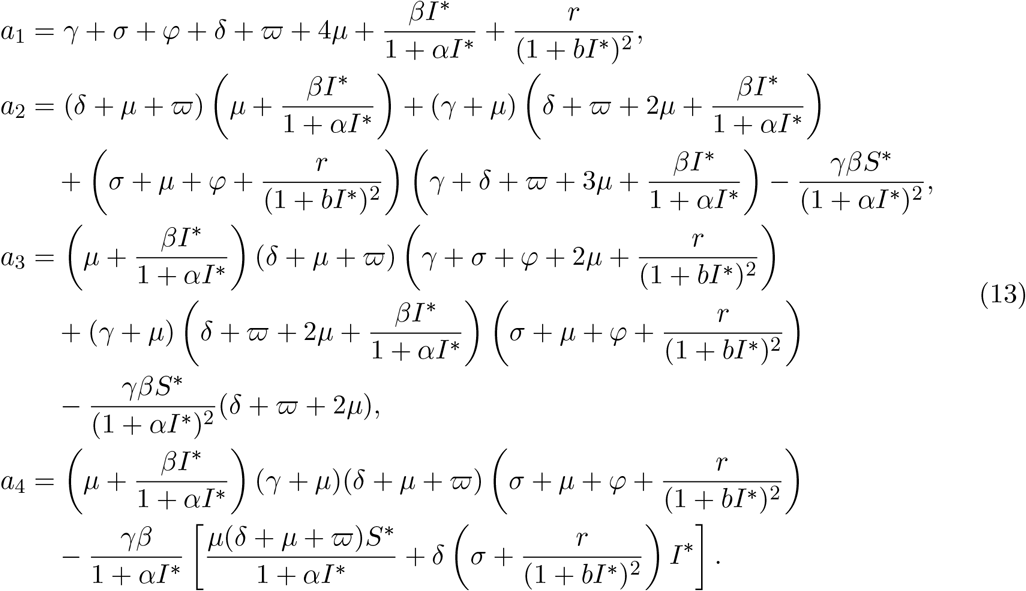

*Proof*. We recall that the Jacobian of system (1) at the endemic state *P*^*\**^ is given by (2). Then, the characteristic equation can be expressed as

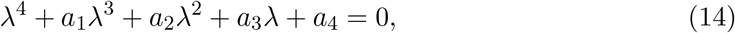

where *a*_1_, *a*_2_, *a*_3_ and *a*_4_ are given in (13).

By the Routh–Hurwitz criterion, it follows that the endemic equilibrium *P*^*\**^ is locally asymptotically stable if and only if *a*_1_ > 0, *a*_3_ > 0, *a*_4_ > 0 and 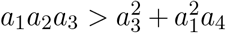. Due to positivity of parameters, *a*_1_ is always positive, so the stability condition can be given by (12). □

## 6 Backward bifurcation analysis

Following the analysis in Section 4, we can obtain a range of values for *R*_0_ in which model (1) can have two positive endemic equilibria. According to Theorem 1, this can only occur when the discriminant 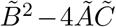 is positive. Thus, to find the value where the two endemic equilibria merge into one, 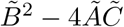 is set to zero and solved for the critical value of *R*_0_, denoted by *R*_*c*_, given by

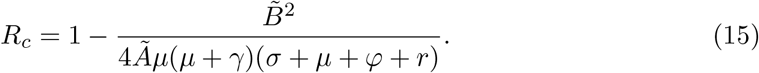

Hence, *R*_*c*_ < *R*_0_ is equivalent to 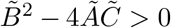 and therefore, backward bifurcation would occur for values of *R*_0_ such that *R*_*c*_ < *R*_0_ < 1. By this and Theorem 1, the following result is established.

### Lemma 1.

*The system* (1) *has two endemic equilibria when* 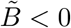 *and R*_*c*_ < *R*_0_ < 1.

At this juncture we establish a stronger result based on a theorem due to Carr [9] and restated in Castillo-Chavez and Song [10], which is based on the general centre manifold theory and is not only useful in determining the local stability of the nonhyperbolic equilibrium but also settles the question of the existence of another equilibrium which has bifurcated from the nonhyperbolic equilibrium.

### Theorem 4.

*Let*

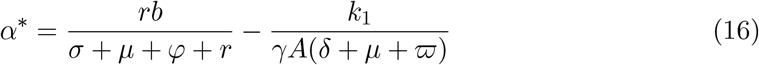

*and*

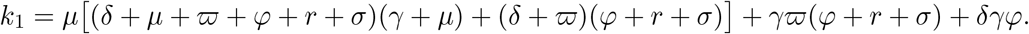

*Then the system of equations in* (1) *undergoes backward bifurcation at R*_0_ = 1 *if α* < *α*^*\**^ *and forward bifurcation if α* > *α*^*\**^

*Proof*. See Appendix B. □

As an example, consider the set of parameters *A* = 1000, *μ* = 0.001, *δ* = 0.2, *γ* = 0.1, *σ* = 1*/*3, *φ* = 0.04, *ϖ* = 0.05, *r* = 1*/*7, *b* = 4 with variable values for *α* and *β*. Then, we can calculate *α*^*\**^ = 1.1047 according to (16). Notice that the value of *R*_0_ varies proportionally to *β*, and *R*_0_ = 1 corresponds to *β* = 5.2236 × 10^−7^. Theorem 4 implies that a backward bifurcation occurs at *R*_0_ = 1 if *α* < 1.1047, while if *α* > 1.1047, the bifurcation is forward.

An example of the bifurcation diagram for model (1) when *α* = 0.1 < *α*^*\**^ can be seen in Figure 2, which depicts the number of infected individuals at equilibria as *R*_0_ varies. There is a critical value *R*_*c*_ = 0.8530 such that for *R*_0_ ∈ (*R*^*\**^, 1) there exist two endemic equilibria. As *R*_0_ crosses the value *R*_*c*_, the two endemic equilibria merge at a limit point (labelled LP in the figure) and disappear via a saddle-node bifurcation.

**Figure 2:**
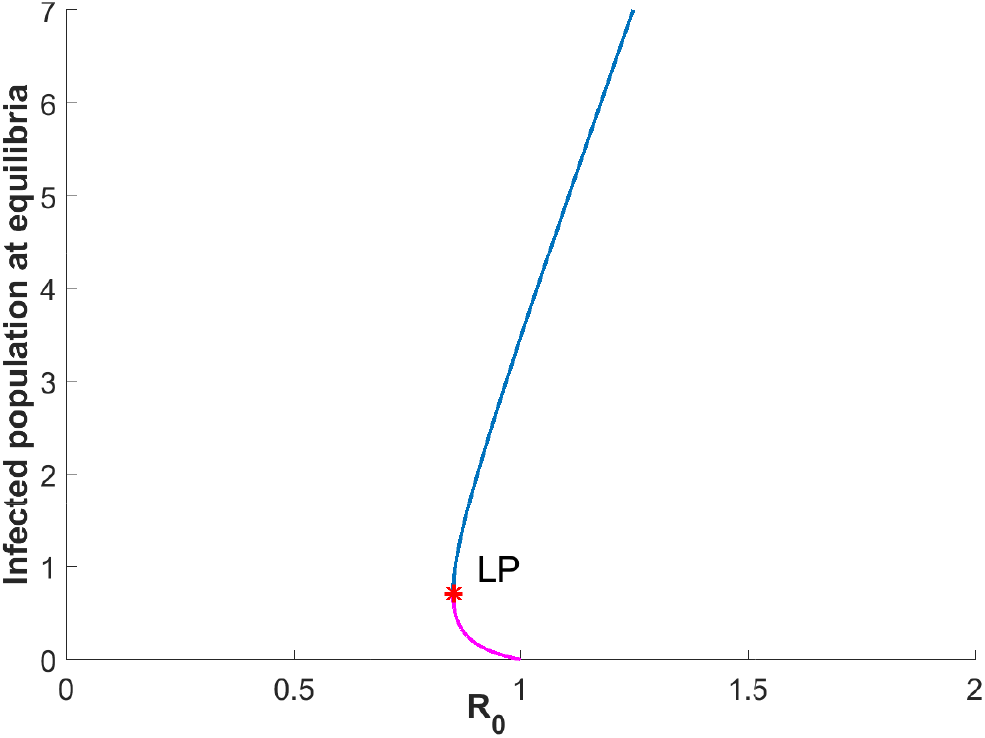
Backward bifurcation diagram of model (1) with parameter values *A* = 1000, *μ* = 0.001, *δ* = 0.2, *γ* = 0.1, *σ* = 1*/*3, *φ* = 0.04, *ϖ* = 0.05, *r* = 1*/*7, *b* = 4 and *α* = 0.1. The horizontal axis represents the value of the basic reproduction number ranging from 0 to 2 as *β* varies from 0 to 1.0447 × 10^−6^. The vertical axis indicates the size of infected population at equilibria. LP denotes the saddle-node bifurcation point, at which the two endemic equilibria merge together.

## 7 Hopf bifurcation analysis

We will now focus on studying the phenomenon of Hopf bifurcation, which could occur around an endemic equilibrium of our system when the real part of two complex, conjugate eigenvalues with nonzero imaginary part crosses zero.

By the analysis in Section 5, we know that the characteristic equation of system (1) at an endemic equilibrium *P*^*\**^(*S*^*\**^, *E*^*\**^, *I*^*\**^, *R*^*\**^) takes the form *λ*^4^ + *a*_1_*λ*^3^ + *a*_2_*λ*^2^ + *a*_3_*λ* + *a*_4_ = 0, where *a*_1_, *a*_2_, *a*_3_ and *a*_4_ are given by (13). Then, following the theory in [7], we can obtain a necessary and sufficient condition for the occurrence of a Hopf bifurcation around *P*^*\**^. This is stated in the following result.

### Theorem 5.

*Let P*^*\**^(*S*^*\**^, *E*^*\**^, *I*^*\**^, *R*^*\**^) *be an endemic equilibrium of* (1). *A Hopf bifurcation generically arises around P*^*\**^ *if and only if*

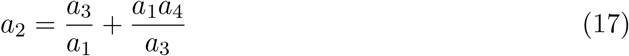

*and* sign(*a*_1_) = sign(*a*_3_).

### 7.1 Numerical example of Hopf bifurcation

For further studying the existence of Hopf bifurcation for system (1), we will choose the treatment rate *r* as a bifurcation parameter. If we substitute the expressions for *a*_1_, *a*_2_, *a*_3_ and *a*_4_ given by (13) in equation (17), we can solve for *r* in the resulting equation and obtain thus a critical value *r* = *r*^*\**^ where the Hopf bifurcation occurs. However, due to the complexity of the calculations, we do not give an explicit expression for the value of *r*^*\**^; instead, we will compute an approximate value using the numerical continuation package Matcont [12], which also allows us to determine the direction of bifurcation.

If we consider the set of parameters *A* = 20289, *μ* = 2.49 × 10^−5^, *σ* = 1*/*10, *γ* = 1*/*4, *δ* = 0.05, *β* = 1.5 × 10^−9^, *α* = 1 × 10^−6^, *b* = 5 × 10^−5^, *φ* = 0.142857, *ϖ* = 0.142857 and *r* = 0.08, we can see that system (1) has a unique endemic equilibrium *P*^*\**^(*S*^*\**^, *E*^*\**^, *I*^*\**^, *R*^*\**^) ≈ (185894582, 70962, 67952, 41639) and *R*_0_ = 3.7850. By an application of Theorem 3, *P*^*\**^ is locally asymptotically stable. Figure 3 shows the solution of system (1) with the initial conditions *S*(0) = 189000000, *E*(0) = 70000, *I*(0) = 60000, *R*(0) = 40000, and it can be seen that all sub-populations converge to positive values, which represent the case when COVID-19 becomes endemic in the population.

**Figure 3:**
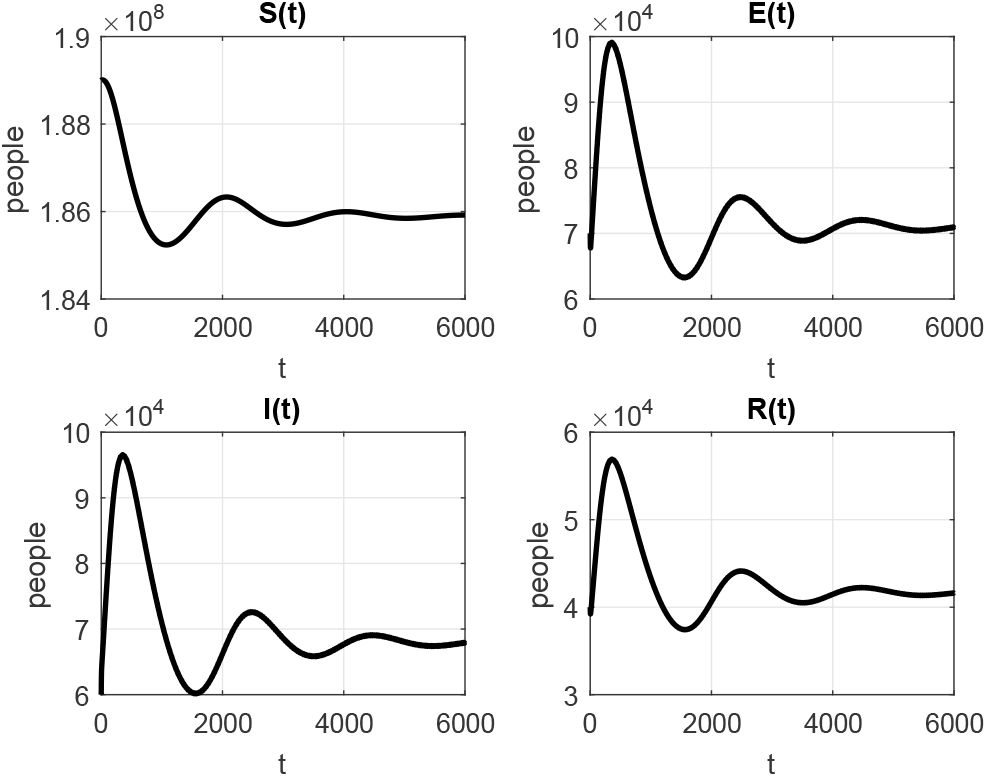
Time evolution of the solutions for model (1) with parameter values *A* = 20289, *µ* = 2.49 × 10^−5^, *σ* = 1*/*10, *γ* = 1*/*4, *δ* = 0.05, *β* = 1.5 × 10^−9^, *α* = 1 × 10^−6^, *b* = 5 × 10^−5^, *φ* = 0.142857, *ϖ* = 0.142857 and *r* = 0.08. In this case, *R*_0_ = 3.7850 and the endemic equilibrium is locally asymptotically stable, which indicates that the disease persists and the infected population approaches a constant level.

However, if we increase the value of the treatment rate *r* while keeping all other parameters fixed, a Hopf bifurcation occurs. Indeed, when *r* = *r*^*\**^: = 0.0948775, the coefficients of the characteristic equation at the unique endemic equilibrium become *a*_1_ = 0.6909, *a*_2_ = 0.09607, *a*_3_ = 7.1989 × 10^−6^ and *a*_4_ = 1.0009 × 10^−6^, which satisfy the hypotheses of Theorem 5. Hence, *P*^*\**^ switches from stable to unstable at *r* = *r*^*\**^. The first Lyapunov coefficient, as computed by Matcont, is 5.11344 × 10^−17^ > 0, which implies that the bifurcation is subcritical. Thus, an unstable limit cycle exists near *P*^*\**^ for values of *r* slightly less than *r*^*\**^.

Figure 4 depicts the solution of the system when *r* = 0.10 > *r*^*\**^ and all other parameters and initial conditions are the same as in Figure 3. In this case, the endemic equilibrium *P*^*\**^(*S*^*\**^, *E*^*\**^, *I*^*\**^, *R*^*\**^) ≈ (189146563, 70805, 66548, 42475) is unstable even though *R*_0_ = 3.5642 is greater than one. We can see in Figure 4 that the number of infected individuals does not settle down at a constant value but presents oscillations that increase in magnitude as time passes.

**Figure 4:**
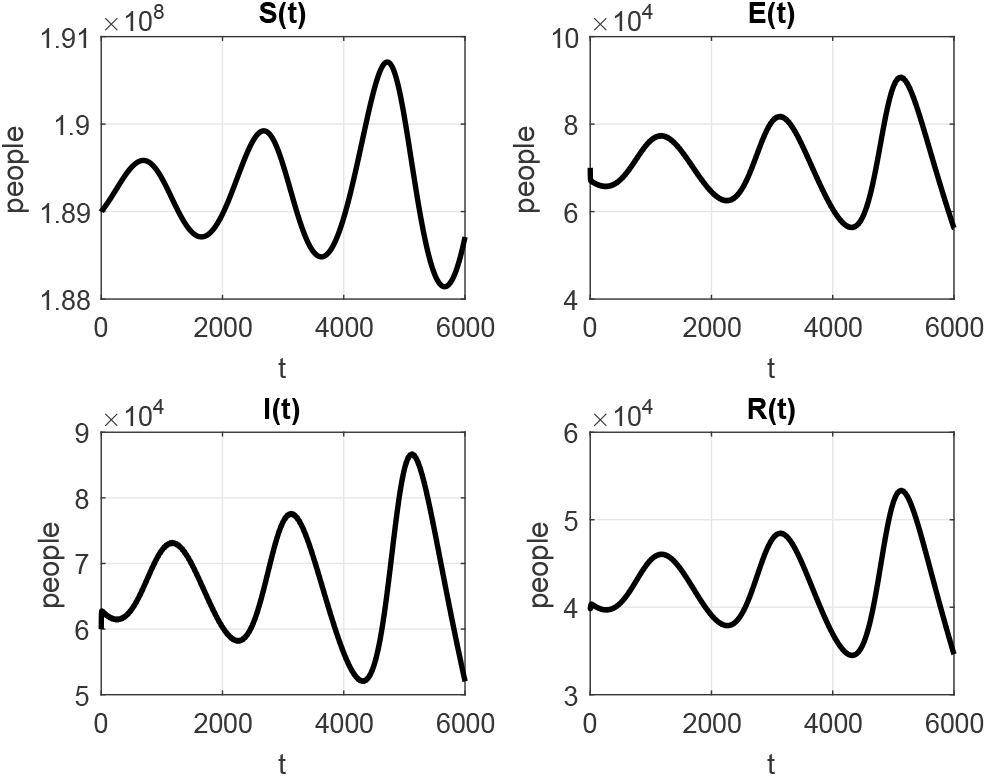
Time evolution of the solutions for for model (1) with parameter values *A* = 20289, *µ* = 2.49 × 10^−5^, *σ* = 1*/*10, *γ* = 1*/*4, *δ* = 0.05, *β* = 1.5 × 10^−9^, *α* = 1 × 10^−6^, *b* = 5 × 10^−5^, *φ* = 0.142857, *ϖ* = 0.142857 and *r* = 0.10. In this case, *R*_0_ = 3.5642 and the endemic equilibrium is unstable. The infection level oscillates around a positive value.

## 8 Parameter fitting based on real data

In this section, we perform some numerical simulations for model (1) using a set of parameter values fitted to the reported data of the COVID-19 pandemic in Nigeria.

Data were collected from the Johns Hopkins University repository [16] for the period from 28 February 2020 to 26 July 2020. We considered the daily data for cumulative infected cases, recovered cases and deaths.

Since it is known that people infected with 2019-nCoV can transmit the pathogen to other people even when they have no visible symptoms of the disease (asymptomatic infection), we need to subdivide the *I*-class of the model into two subclasses: symptomatic infectious (*I*_1_(*t*)) and asymptomatic (*I*_2_(*t*)) infectious. Individuals in the exposed population become infectious at a rate *γ*. This means that, after 1*/γ* time units, an exposed individual becomes symptomatically infectious with a probability *p* or asymptomatically infectious with a probability 1 − *p*. Hence, for performing the parameter fitting and simulations, we consider the system

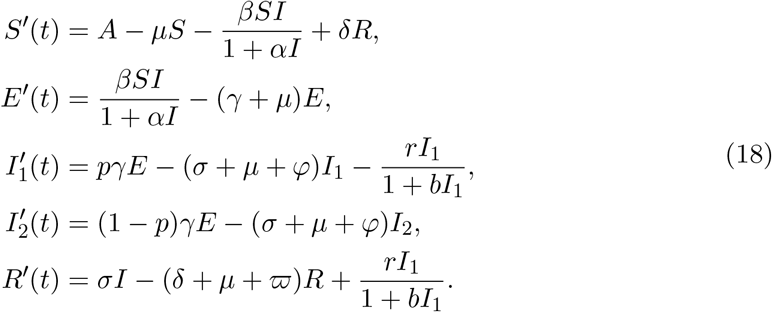

Notice that the saturated treatment response *T* (*I*) = *rI/*(1+*bI*) is included in the equation for *I*_1_ but not in the equation for *I*_2_, because asymptomatic infectious individuals recover naturally from the disease without receiving treatment.

In order to compare the temporal dynamics of each class as predicted by the model with the real data, we will also consider the cumulative number of symptomatic infectious cases, which we will denote by *C*(*t*) and is governed by the equation

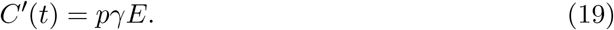

We denote by *R*_1_(*t*) the number of individuals removed by death due to the virus. The dynamics of this class can be described by

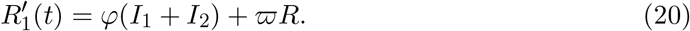

The parameter values used in the simulations are shown in Table 1. The values for the recruitment rate *A* and the natural death rate *μ* were chosen according to the birth and death statistics for the population of Nigeria [11]. The values for *σ, p* and *γ* are biological constants that have been estimated in the literature [19, 23, 26]. The rate of transfer *δ* from the recovered population to the susceptible population is considered variable, and the rest of the parameter values were fitted according to the reported data of cumulative infected cases, recovered cases and deaths.

**Table 1:** Values of the parameters used in numerical simulations.

| Parameter | Value | Unit | Source |
| --- | --- | --- | --- |
| $A$ | 20289 | people/day | [11] |
| $\mu$ | $2.49 \times 10^{-5}$ | 1/day | [11] |
| $\sigma$ | 1/10 | 1/day | [19] |
| $p$ | 0.7 | – | [23] |
| $\gamma$ | 1/4 | 1/day | [26] |
| $\delta$ | Variable | 1/day | Assumed |
| $\beta$ | $1.5 \times 10^{-9}$ | 1/(people · day) | Estimated |
| $\alpha$ | $3.33 \times 10^{-4}$ | 1/people | Estimated |
| $r$ | $1 \times 10^{-4}$ | 1/day | Estimated |
| $b$ | $5 \times 10^{-5}$ | 1/people | Estimated |
| $\varphi$ | $1 \times 10^{-5}$ | 1/day | Estimated |
| $\varpi$ | $5 \times 10^{-4}$ | 1/day | Estimated |

We considered first the case of no reinfection, that is, when *δ* = 0. In this case, a comparison of the estimated dynamics of the model and the reported data can be seen in Figure 5 for the cumulative symptomatic infections, Figure 6 for the recovered cases and Figure 7 for the number of deaths. We also plot the estimated number of exposed individuals in Figure 8 and active infectious individuals (symptomatic and asymptomatic) in Figure 9.

**Figure 5:**
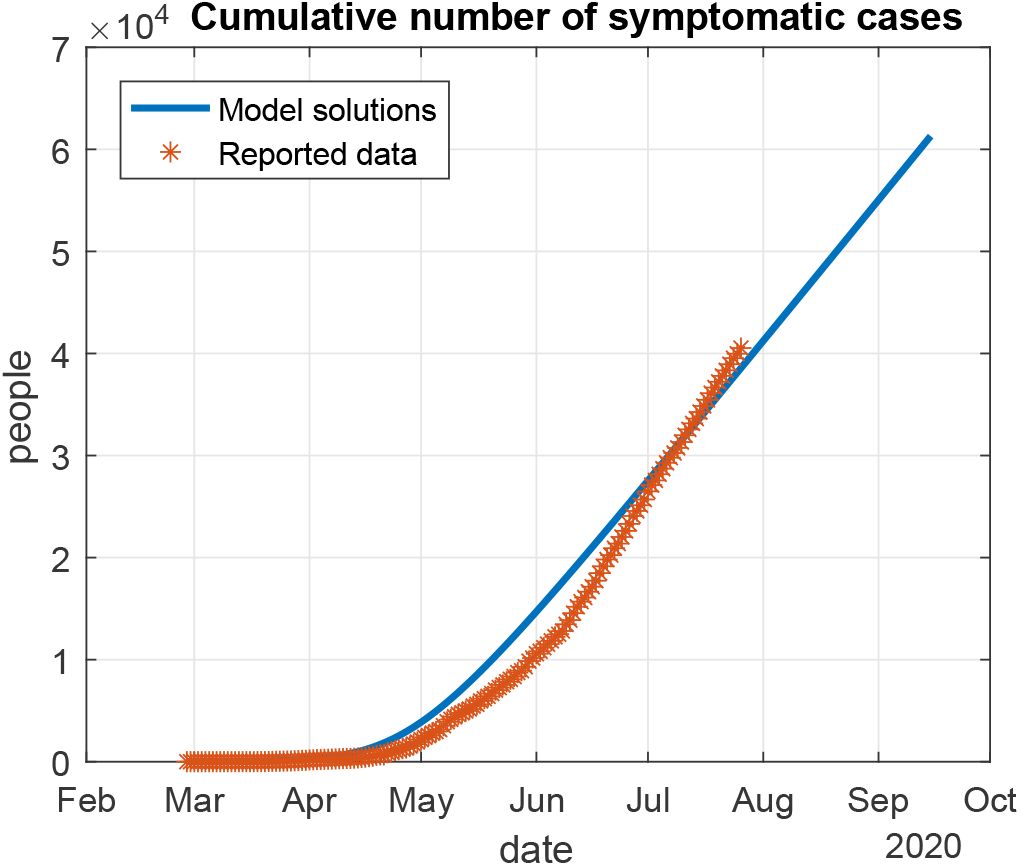
Cumulative number of symptomatic infectious individuals estimated by the model (*C*(*t*)) and reported data. The solutions of model (18) were computed using the parameter values from Table 1 and *δ* = 0. Data shown is the reported number of cumulative infected cases in Nigeria.

**Figure 6:**
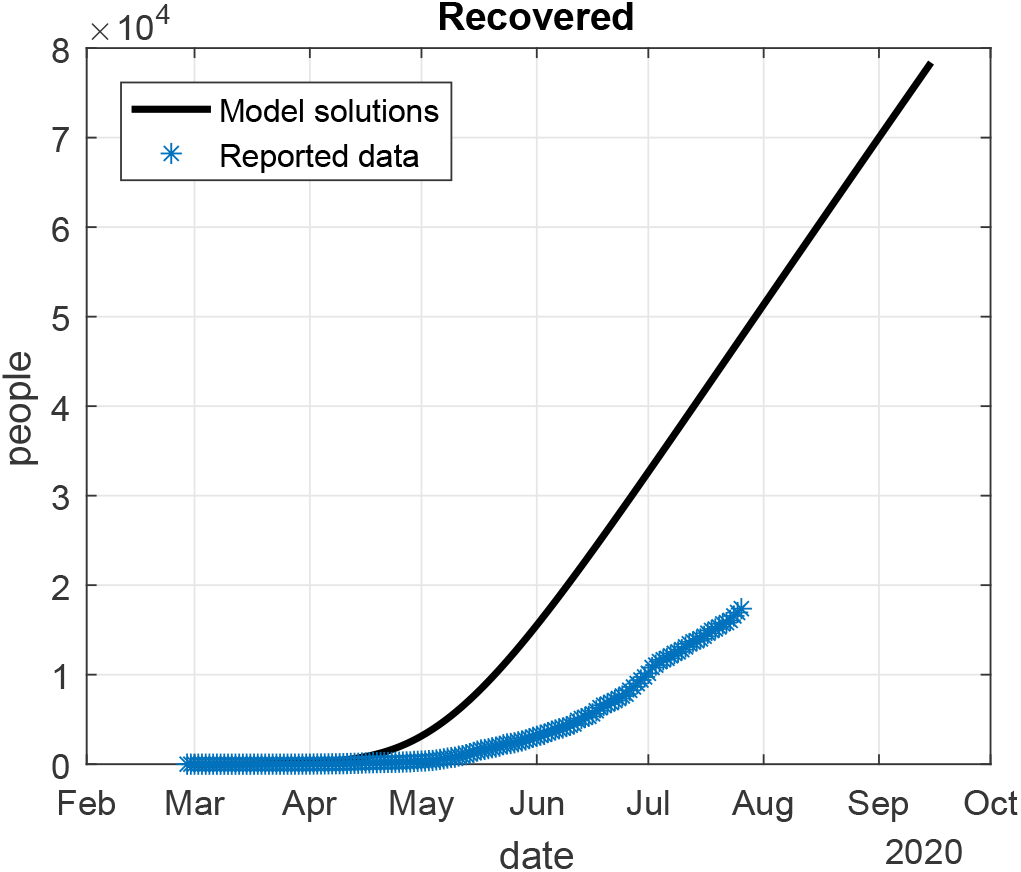
Number of recovered individuals estimated by the model (*R*(*t*)) and reported data. The solutions of model (18) were computed using the parameter values from Table 1 and *δ* = 0. Data shown is the reported number of recovered cases in Nigeria.

**Figure 7:**
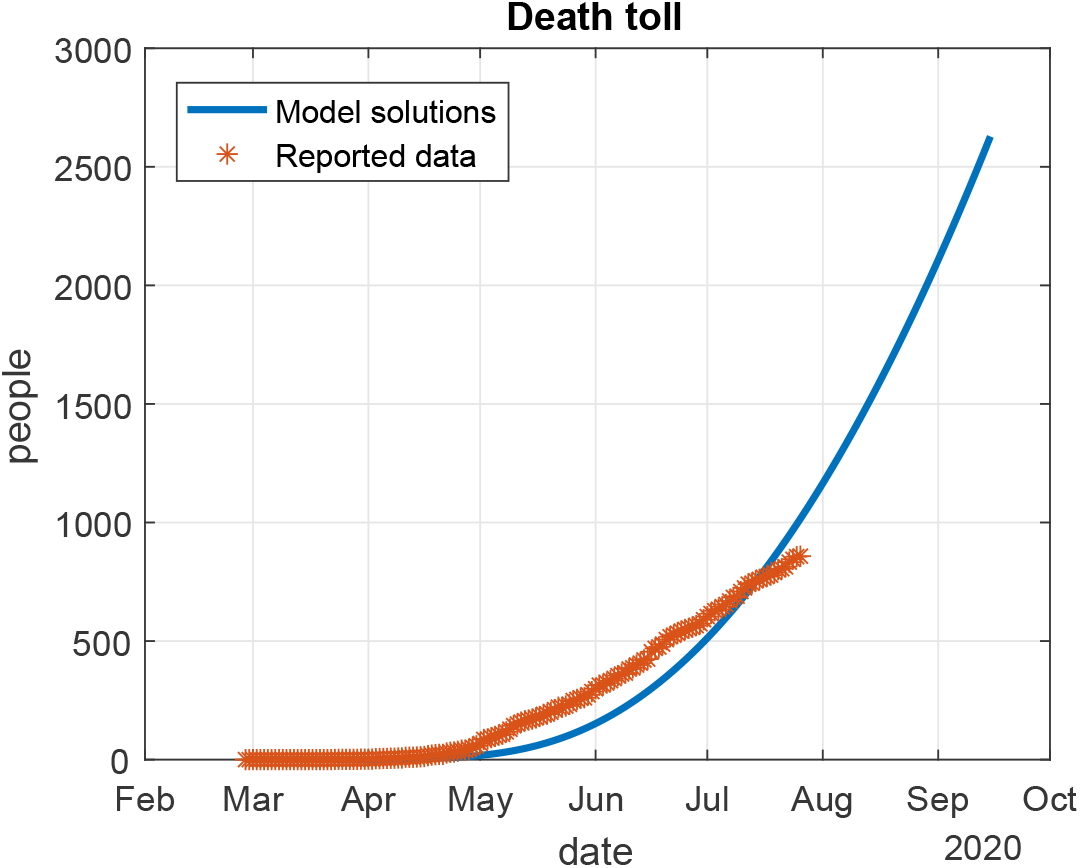
Death toll estimated by the model (*R*_1_(*t*)) and reported data. The solutions of model (18) were computed using the parameter values from Table 1 and *δ* = 0. Data shown is the reported number of COVID-19 deaths in Nigeria.

**Figure 8:**
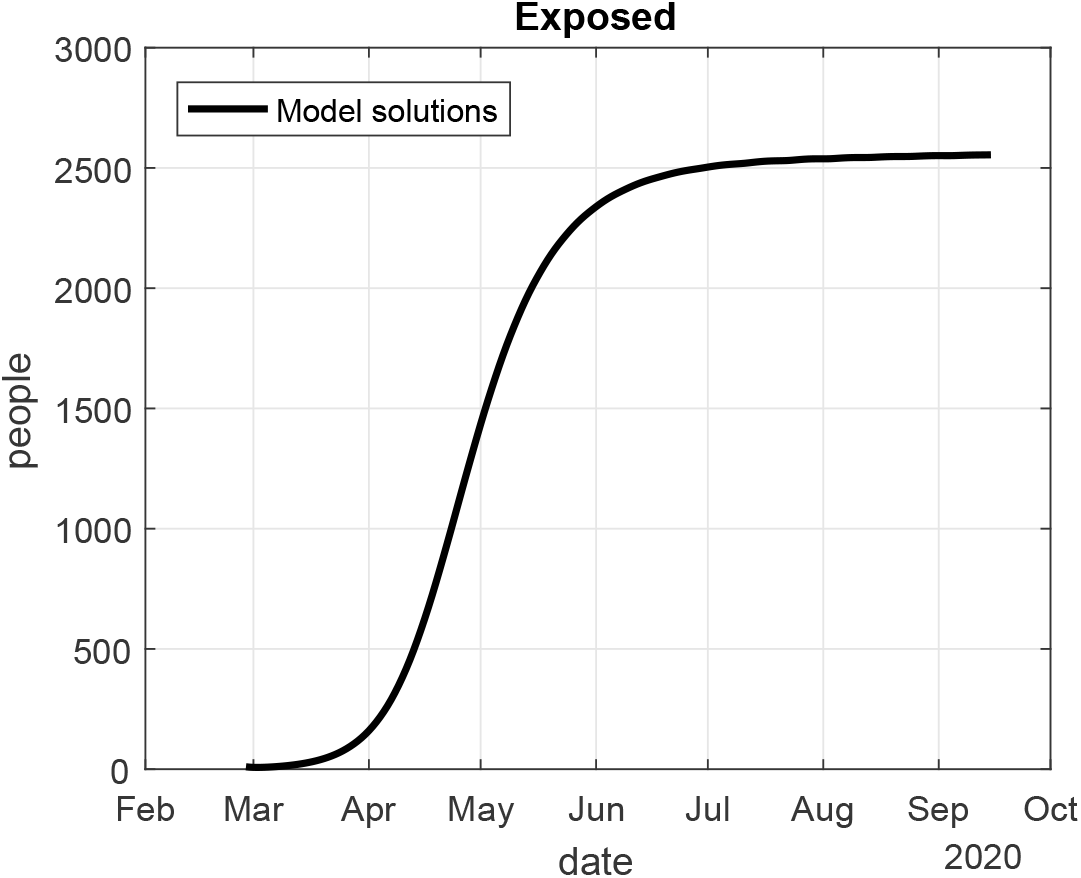
Number of exposed individuals estimated by the model (*E*(*t*)). The solutions of model (18) were computed using the parameter values from Table 1 and *δ* = 0.

**Figure 9:**
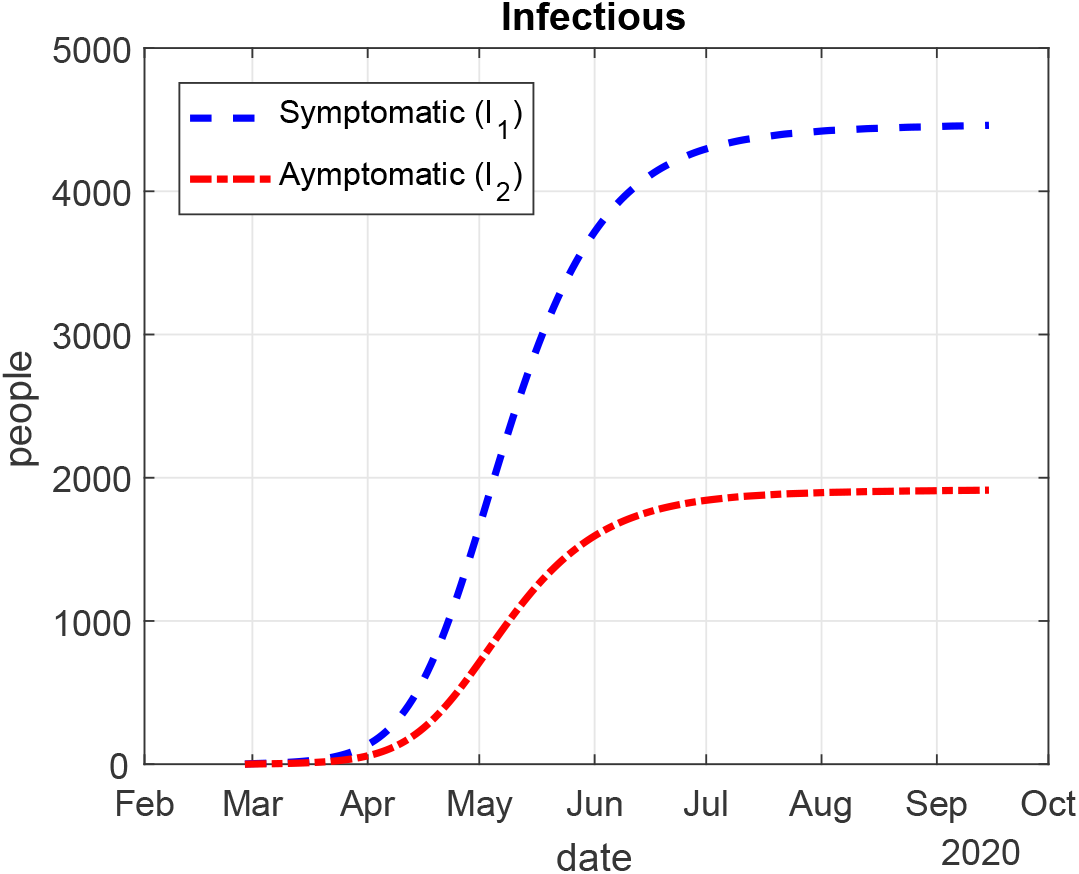
Number of symptomatic (*I*_1_(*t*)) and asymptomatic (*I*_2_(*t*)) infectious individuals estimated by the model. The solutions of model (18) were computed using the parameter values from Table 1 and *δ* = 0.

Our simulations indicate that the cumulative number of infected and recovered cases will keep on increasing in an almost linear fashion, at least until October 2020. The number of deaths will also increase, but its growth rate will become larger as time passes (see Figure 7). This implies that there will still be a long time until the pandemic goes extinct. Although the official reported data do not include information on the number of exposed individuals, our simulations show that the exposed cases would stabilize at a number slightly larger than 2500 people from July 2020 onwards (Figure 8).

In the case when reinfection is possible, the value 1*/δ* represents the average time that an individual spends in the recovered class before becoming susceptible to a reinfection by 2019-nCoV, i.e., 1*/δ* is the length of the immunity period. We plotted the dynamics of the model for several values of *δ*, including the case *δ* = 0 (permanent immunity): Figure 10 shows the recovered cases and Figure 11 represents the death toll. The graphs for the other compartments are not shown since there are no perceivable differences with the graphs for the case *δ* = 0.

**Figure 10:**
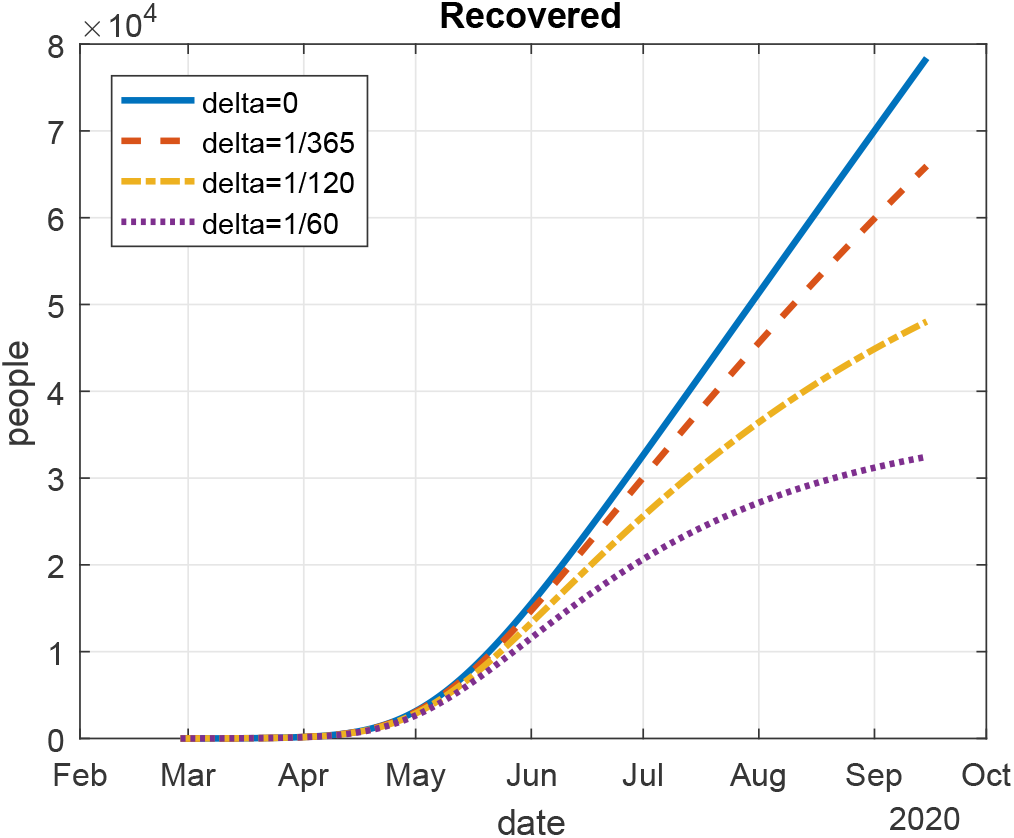
Number of recovered individuals (*R*(*t*)) estimated using the solutions of model (18) for different values of *δ*. The values for other parameters were taken as in Table 1. The simulations show that the value of *R*(*t*) is decreasing as the length of the immunity period decreases.

**Figure 11:**
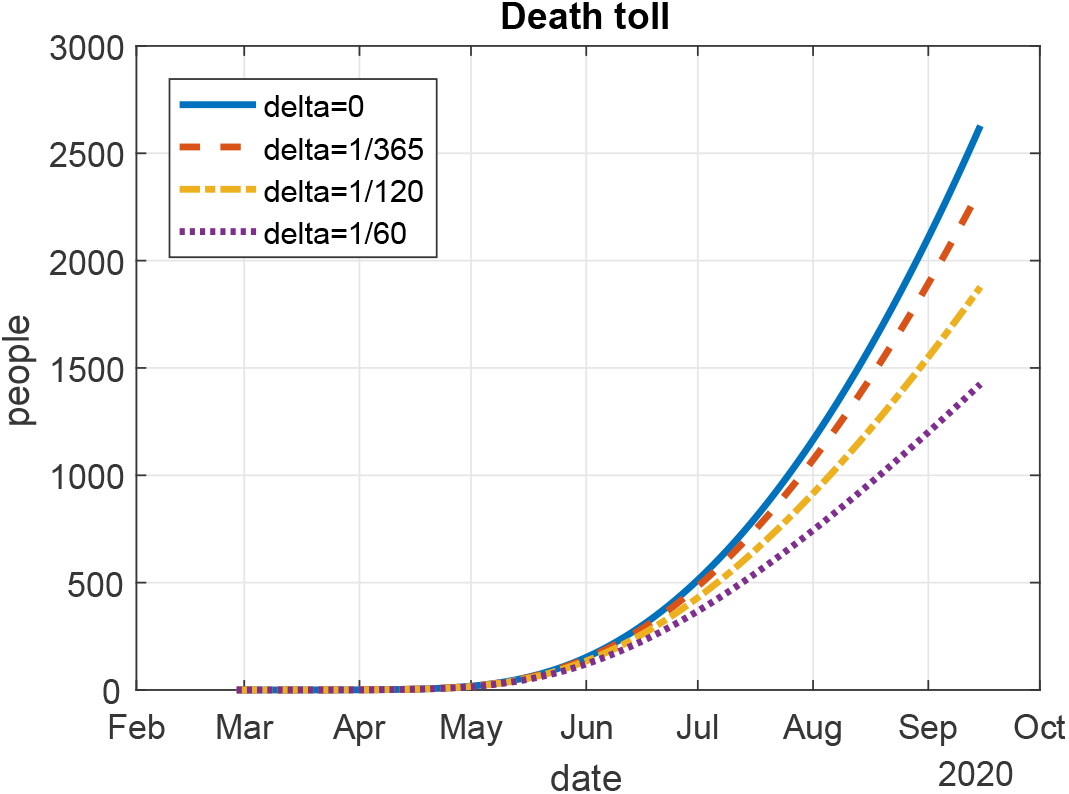
Death toll (*R*_1_(*t*)) estimated using the solutions of model (18) for different values of *δ*. The values for other parameters were taken as in Table 1. The simulations show that the value of *R*_1_(*t*) is decreasing as the length of the immunity period decreases.

Our results show that the number of recoveries and deaths would be lower in the case when reinfection is possible after recovery; moreover, it becomes even lower as the length of the immunity period is reduced from one year to 60 days. On the other hand, the cumulative number of infections is not visibly affected by the possibility of reinfection, at least for the time period we considered in our simulations.

## 9 Discussion and conclusion

We proposed and studied an SEIRS COVID-19 epidemic model that includes saturated incidence and a saturated Holling type II treatment response. Due to the limitations in the number of hospital beds and intensive care units that exist in every country, we believe that this model is more realistic than those with a linear treatment response, which grows at the same rate irrespectively of the number of infected people at a given time. We have extended the results published in [2], where a similar model was studied for a general disease, but the authors there did not delve into the study of bifurcation dynamics. In the present paper, we discussed the backward and Hopf bifurcation of model (1) to help government and policy makers decide an efficient response plan to combat a second wave of the COVID-19 pandemic, which has been widely reported in places like Asia and Europe.

Epidemiologically, we understand the role of reproduction number in controlling disease, but there are times that *R*_0_ < 1 does not represent the eradication of the disease and at that critical phase a reemergence can occur which may be more endemic than the first. Our analysis showed that model (1) presents the phenomenon of backward bifurcation for certain values of the parameters. When this type of bifurcation occurs, the eradication of the epidemic may not be guaranteed by simply reducing the basic reproduction number *R*_0_ below unity; instead, *R*_0_ should be further reduced to a critical value *R*_*c*_ < 1.

We performed some numerical simulations for model (1) using a set of parameter values fitted to the reported data of the COVID-19 pandemic in Nigeria collected from the Johns Hopkins University [16] between 28 February 2020 and 26 July 2020. We considered the daily data for cumulative infected cases, recovered cases and deaths. As we proved in Section 4, backward bifurcation cannot take place if the parameter *b*, which measures the saturation effect in treatment response, is sufficiently close to 0. The reduction of this parameter, however, can only be achieved when a community has sufficient medical capacity for the treatment of COVID-19, which is an unrealistic assumption. A more plausible way to avoid the backward bifurcation scenario is by increasing the parameter *α* so that it becomes larger than *α*^*\**^ (see Theorem 4). A larger value for *α* indicates that the incidence function *βSI/*(1 + *αI*) saturates for smaller values of *I*. The interpretation of this is that the number of infectious contacts between persons must be reduced at a time when the number of infected individuals is still small. Therefore, we conclude that the application of social distancing and stay-at-home policies should be made since the early stages of the epidemic.

Although backward bifurcation is not a new phenomenon in the study of epidemic disease dynamics, several possible causes for the appearance of this bifurcation in COVID-19 models have been identified. For example, the authors in [22] studied a model with a compartment for lockdown population and analysed the effects of lockdown efficacy; they showed that the backward bifurcation can be caused by an imperfect lockdown. In [17], the authors studied a COVID-19 model that can present backward bifurcation only if the recovered individuals have temporary immunity, instead of permanent immunity. We should remark, however, that the models in [22] and [17] did not include saturation effects in the treatment rate. Our results for model (1) show that the limitation of medical resources alone can be a cause for bistability behaviour and backward bifurcation, similarly to what was studied in [21] and [3]. Moreover, we considered a saturation effect in the incidence rate that was not included in the above mentioned literature.

We proved that the inclusion of both saturated incidence and saturated treatment is also a cause of Hopf bifurcation. This is a topic of research that has been scarcely studied for the dynamics of COVID-19 (see, e.g., the analysis of delay-induced Hopf bifurcation in [1] and [25]) and has important implications for epidemic control, because this type of bifurcation can produce oscillatory patterns in the number of infected individuals. Further research should still be made to improve our understanding of the different dynamics that can occur in this epidemic outbreak.

## Data Availability

The data used in this manuscript were publicly available data retrieved from Johns Hopkins University Repository.

https://github.com/CSSEGISandData/COVID-19

## Appendix A Proof of Theorem 1

We will determine the endemic equilibria of the model by considering equations (5)–(8) when *I* is positive. Solving for *E* and *R* in equations (7) and (8), respectively, we obtain

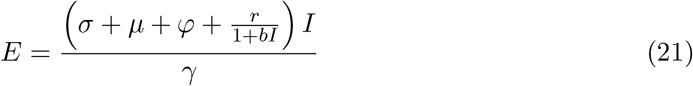

and

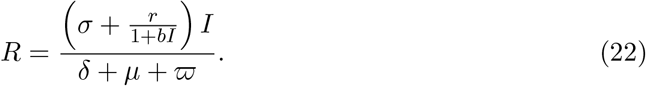

Then, from equations (6) and (21), we also get

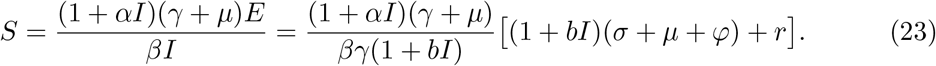

Lastly, it follows from (5) and (6) that *A*−*μS* + *δR*− (*γ* + *μ*)*E* = 0. Substituting equations (21)–(23) and simplifying, we obtain the following equation in *I*:

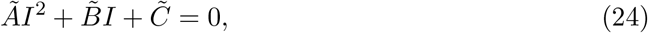

where 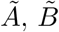 and 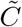 are defined in (9).

Therefore, the endemic equilibria of system (1) take the form (*S*^*\**^, *E*^*\**^, *I*^*\**^, *R*^*\**^), where

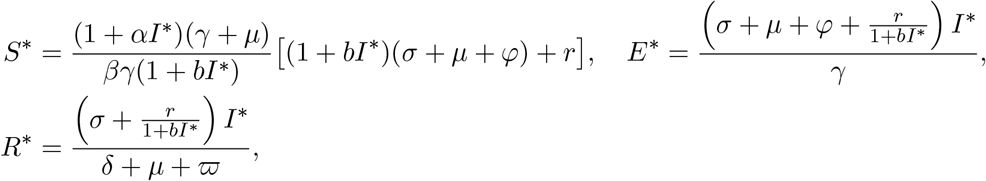

and *I*^*\**^ is a positive root of the polynomial

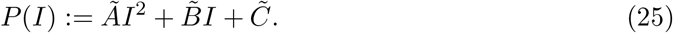

Since *δ* ≥ 0 and the other parameters of the model are positive, we have 0 ≤ *δ/*(*δ* +*μ*+ *ϖ*) < 1, and then

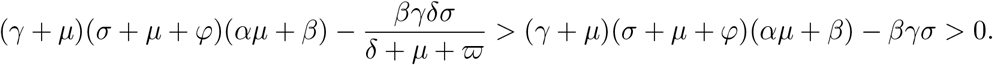

Thus, the coefficient *Ã* in (9) is always positive, and 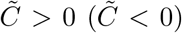 if *R*_0_ < 1 (*R*_0_ > 1, respectively). Since *Ã* > 0, the existence of the positive solutions of (25) depends on the signs of 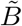 and 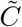. If *R*_0_ > 1, then 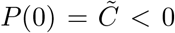 and the graph of *P* (*I*) is an upwards parabola that crosses the horizontal axis at a positive value and a negative value of *I* (see Figure 12); hence, (25) has exactly one positive root and thus there is a unique endemic equilibrium.

**Figure 12:**
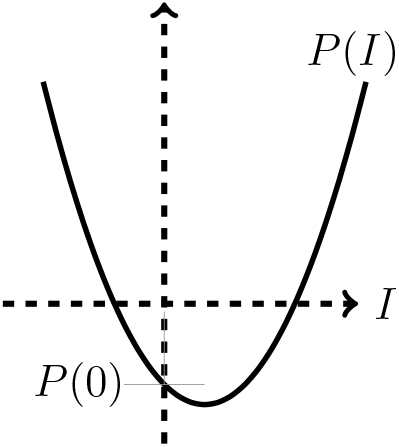
Graph of *P* (*I*) in the case when *R*_0_ > 1.

If *R*_0_ = 1, then 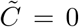 and there is a unique non-zero solution of (25), 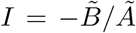, which is positive if and only is 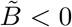. In this case, there is a positive endemic equilibrium if 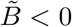, and none otherwise.

Lastly, we consider the case when *R*_0_ < 1 and then 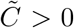. If 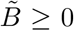, it is easy to see by Descartes’ rule of signs that *P* (*I*) has no positive roots. If 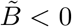, the solutions of (25) are given by

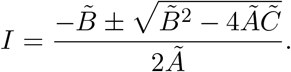

These solutions are positive and distinct only when 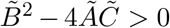, in which case there are two endemic equilibria. The solutions of (25) coalesce into a double root when 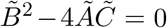, so there is only one endemic equilibrium in that case.

## Appendix B Proof of Theorem 4

We simplify and change the variables on the system (1). Let *S* = *x*_1_, *E* = *x*_2_, *I* = *x*_3_ and *R* = *x*_4_, so that *N* = *x*_1_ + *x*_2_ + *x*_3_ + *x*_4_. Using vector notation *X* = (*x*_1_, *x*_2_, *x*_3_, *x*_4_)^*T*^, system (1) can be written in the form

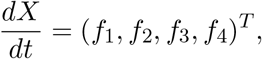

where

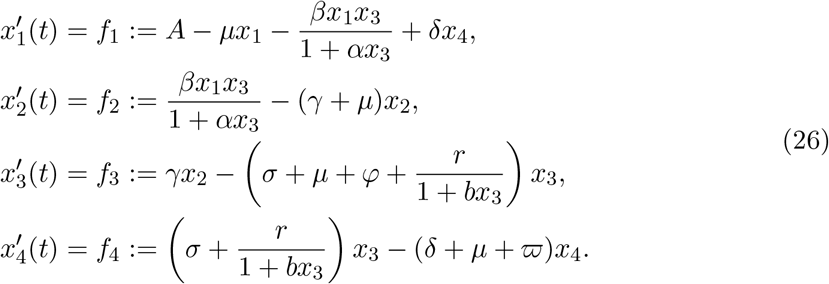

Consider the case when *R*_0_ = 1. Suppose that *β* is chosen as a bifurcation parameter. Solving for *β* from *R*_0_ = 1 gives

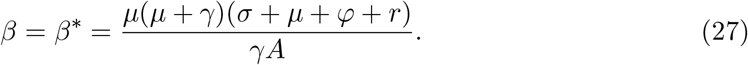

The eigenvalues of Jacobian of the system (1), evaluated at *P*_0_ with *β* = *β*^*\**^, are given by

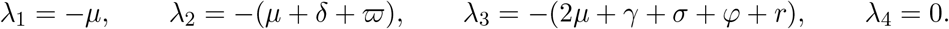

Thus *λ*_4_ = 0 is a simple zero eigenvalue and the other eigenvalues are real and negative. Hence, when *β* = *β*^*\**^ (equivalently, when *R*_0_ = 1), the disease-free equilibrium *P*_0_ is a non-hyperbolic equilibrium, the assumption (A1) in Theorem 4.1 of [10] is thus verified on the system (1). Hence, a right eigenvector associated with the zero eigenvalue *λ*_4_ = 0 is given by

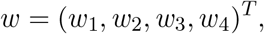

where

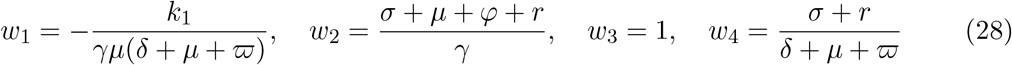

and

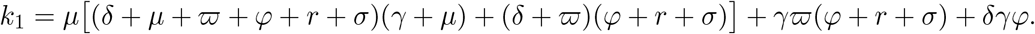

Furthermore, we obtain a left eigenvector associated with the zero eigenvalue *λ*_4_ = 0 satisfying *v* · *w* = 1 given by

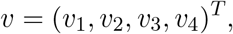

where

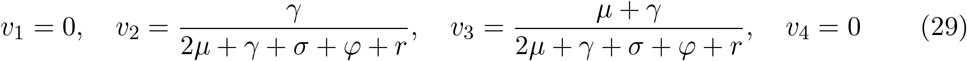

The coefficients *ã* and 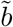 defined by Theorem 4.1 in [10] are computed as follows.

Considering the system of equations in (26), the associated non-zero second partial derivatives of the right-hand side functions (*f*_*i*_) evaluated at (*P*_0_, *β*^*\**^) are given by

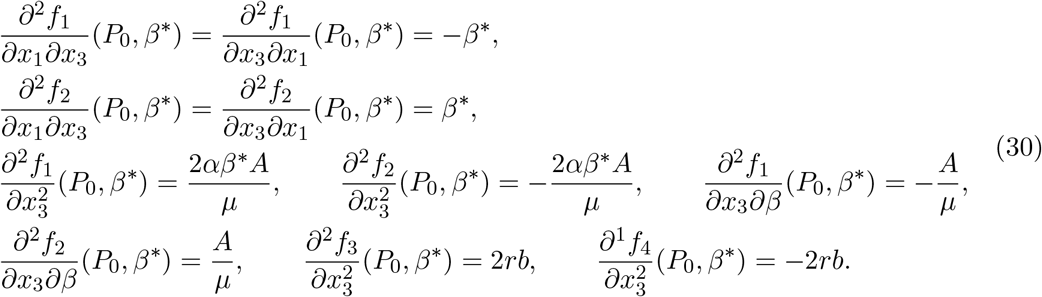

Thus, using the expressions (27)–(30), we compute ã and 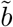 as

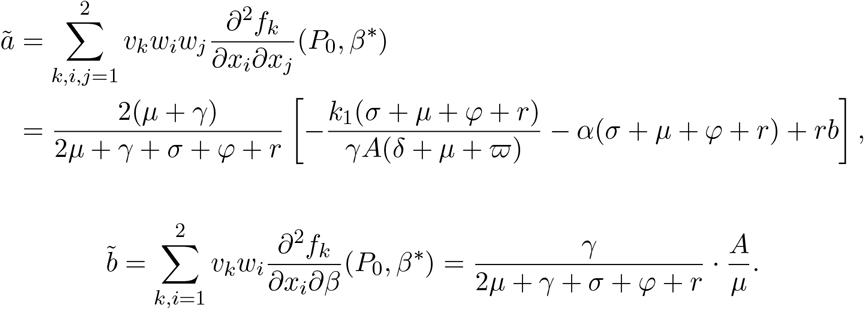

It is found that the coefficient 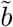 is always positive. The coefficient *a* is positive if *α* < *α*^*\**^ and negative if *α* > *α*^*\**^, where *α*^*\**^ is given by (16). Therefore, by [10, Theorem 4.1], system(1) undergoes backward bifurcation if *α* < *α*^*\**^ and forward bifurcation if *α* > *α*^*\**^

## Notes

### Competing Interest Statement

The authors have declared no competing interest.

### Funding Statement

No external funding was received for this work.

### Author Declarations

IRB approval was not required since the work used only publicly available data.

### Summary of Updates

Proofs of Theorems 1 and 4 were moved to an appendix; figure captions were updated

